# Early introduction and rise of the Omicron SARS-CoV-2 variant in highly vaccinated university populations

**DOI:** 10.1101/2022.01.27.22269787

**Authors:** Brittany A. Petros, Jacquelyn Turcinovic, Nicole L. Welch, Laura F. White, Eric D. Kolaczyk, Matthew R. Bauer, Michael Cleary, Sabrina T. Dobbins, Lynn Doucette-Stamm, Mitch Gore, Parvathy Nair, Tien G. Nguyen, Scott Rose, Bradford P. Taylor, Daniel Tsang, Erik Wendlandt, Michele Hope, Judy T. Platt, Karen R. Jacobson, Tara Bouton, Seyho Yune, Jared R. Auclair, Lena Landaverde, Catherine M. Klapperich, Davidson H. Hamer, William P. Hanage, Bronwyn L. MacInnis, Pardis C. Sabeti, John H. Connor, Michael Springer

## Abstract

The Omicron variant of SARS-CoV-2 is transmissible in vaccinated and unvaccinated populations. Here, we describe the rapid dominance of Omicron following its introduction to three Massachusetts universities with asymptomatic surveillance programs. We find that Omicron was established and reached fixation earlier on these campuses than in Massachusetts or New England as a whole, rapidly outcompeting Delta despite its association with lower viral loads. These findings highlight the transmissibility of Omicron and its propensity to fixate in small populations, as well as the ability of robust asymptomatic surveillance programs to offer early insights into the dynamics of pathogen arrival and spread.

## Introduction

In the final days of 2021, the global SARS-CoV-2 case count surpassed 1 million confirmed cases per day^1^, with the surge due at least in part to the Omicron variant of concern (B.1.1.529). In the United States, COVID-19 case counts reached record highs (3-5 times the peak of prior waves), with the estimated percentage of cases due to Omicron rapidly increasing from < 1% of cases (December 4) to > 95% of cases (January 1)^2^. Omicron transmission is clearly possible among both vaccinated and unvaccinated individuals^3^, although the relative rates in each remain unclear. Evidence suggests that Omicron can at least partially evade immunity acquired from prior COVID-19 infection^4^ and from a two-dose mRNA vaccine regimen^5^, though a third mRNA vaccine dose improves Omicron neutralization efficiency^6^.

To mitigate the risks of congregate living, institutes of higher education (IHEs) use a combination of vaccination requirements^7,8^, high-frequency testing^8,9^, and behavioral interventions such as masking and social distancing to control viral spread on campuses. An analysis^10^ suggests that in the setting of masking and frequent testing, case counts are not correlated with dorm occupancy or in-person instruction; this is consistent with the evidence that cases have been predominantly acquired in off-campus settings^11^. Moreover, detailed genomic analyses of an IHE and its nearby communities suggested that transmission dynamics within the IHE did not necessarily result in spread to the greater community^12^. Thus, many IHEs successfully controlled the spread of COVID-19 up through the Delta surge. However, in December 2021, COVID-19 case counts rose rapidly both in college communities^13^ and in New England as a whole, with viral genomic sequencing confirming Omicron as the cause. While some institutions responded by converting to distance learning or requiring booster shots,^14,15,16,17,18^ the feasibility of maintaining residential college life without another spike in cases remains uncertain.

In this work, we describe the dynamics of Omicron at three IHEs – Boston University (BU), Harvard University (HU), and Northeastern University (NU) – in the greater Boston area. We document the rise of the Omicron variant relative to the Delta (B.1.617.2) variant, and its rapid approach to fixation. We show that the establishment of Omicron at IHEs precedes that of the state and region, and that the time to fixation is shorter at IHEs than in the state or region. We show that university employees have an Omicron trajectory that resembles that of students, with a 2-3 day delay. Finally, we compare cycle threshold (Ct) values in Omicron *vs*. Delta variant cases on college campuses, and identify lower viral loads among college affiliates harboring Omicron infections. In summary, we capitalize on asymptomatic testing programs at IHEs to document the rapid takeover of the Omicron variant, reaching near-fixation within the span of 8-13 days despite lower viral loads, on average, than the previously dominant Delta variant.

## Methods

### Patient samples & ethics statement

We gathered de-identified sample information from three institutions with campus testing programs^11^ (**Table 1**). We received the following information for every positive test collected between December 2 and December 21, 2021: sample collection date, cycle threshold (Ct) for one or more genes, and variant designation. From BU, we also received affiliate status (student *vs*. employee, where employees include faculty, staff, and contractual employees). For HU, SARS-CoV-2 samples were collected from consented individuals under Harvard Longwood Campus IRB #20-1877 and covered by an exempt determination (EX-7295) at the Broad Institute of MIT and Harvard. For BU, SARS-CoV-2 samples and data access were covered by an exemption determination under Boston University IRB #6122E. The use of these data in this study was evaluated and approved by NU under Data Use Agreement 20-1481 and was covered by an exempt determination via Northeastern University IRB #21-02-07.

**Table 1.**
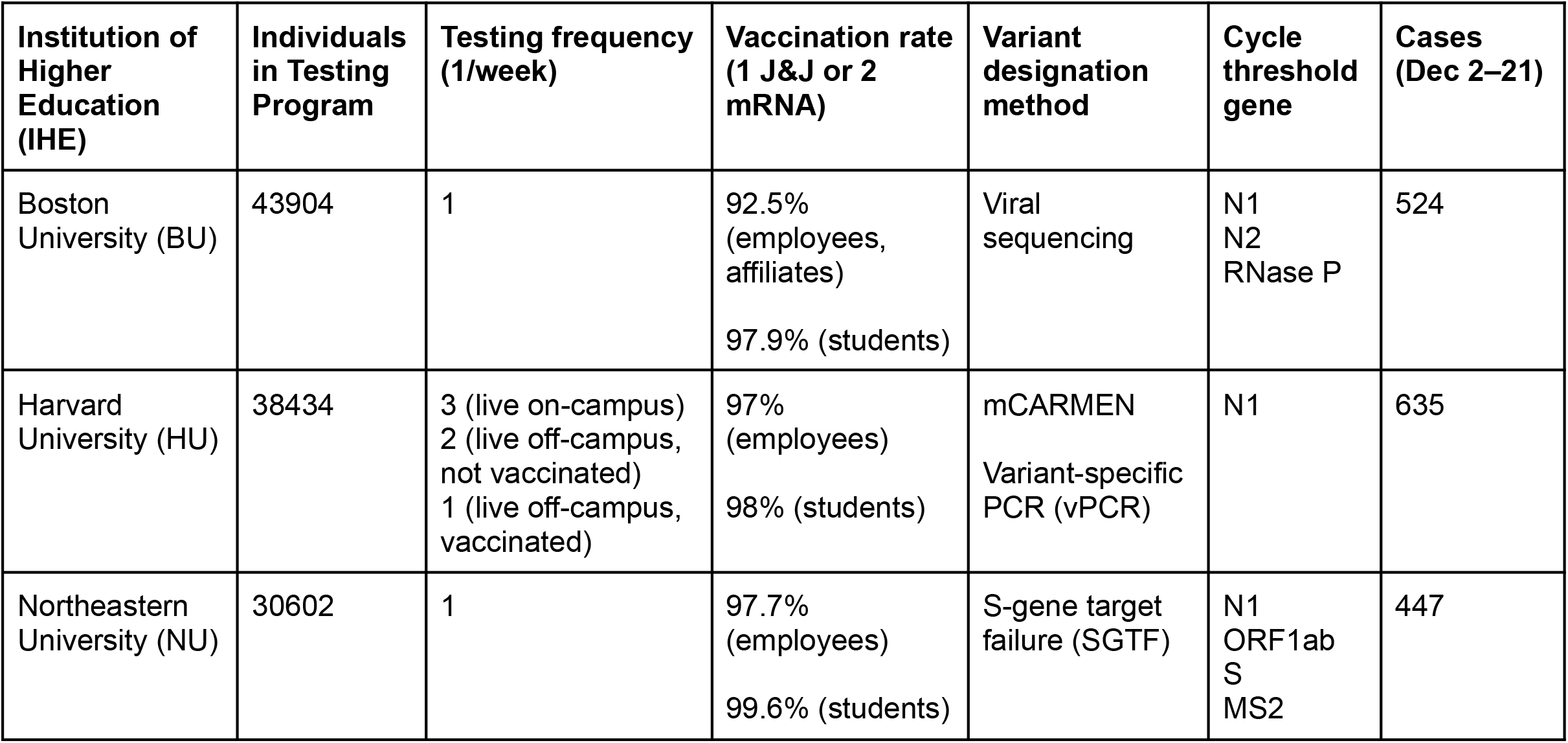
Institutions studied in this work. Individuals in the testing programs include undergraduate and graduate students, faculty, and staff. Testing frequencies and vaccination rates were collected from publicly available university dashboards^45–47^. J&J = Johnson & Johnson vaccine. N1, N2, ORF1ab, and S are SARS-CoV-2 genes; RNase P is a human gene (control); MS2 is a bacteriophage (control).

### Experimental methods

Across universities, individuals self-collected anterior nares specimens, which were analyzed by RT-qPCR. At BU, a two-target SARS-CoV-2 assay with RNase P control was performed^11,19^. At HU, the Quaeris SARS-CoV-2 assay was performed^20^. At NU, the Thermo Fisher Scientific Applied Biosystems™ TaqPath™ COVID-19 Combo Kit was performed^21^. Variant status was assessed using (1) amplicon-based viral sequencing, as previously described^22^ (BU); (2) mCARMEN^23^, a multiplexed CRISPR-based diagnostic platform that identifies unique combinations of Spike gene mutations (HU); (3) a variant-specific PCR assay (HU; **Supplementary Table 1**); and (4) S-gene target failure^24^ (SGTF; NU; **Supplemental Methods**).

## Analytic methods

### Data curation

We downloaded MA and New England (NE) case count data from the CDC^25^. We removed 152 of 1,758 samples (8.6%) from the universities (50 from BU and 102 from HU) with missing variant information (*i*.*e*., due to assay technical limitations) from all subsequent analyses. We removed 53 of the 22,211 (0.2%) MA sequences from GISAID^26–28^ that had a variant classification other than Delta or Omicron. We removed 22,211 of the 30,796 (72.1%) NE sequences from GISAID^26–28^ that were in MA (*i*.*e*., NE curve fits do not include MA), and 29 of the remaining 8,585 (0.3%) sequences that had a variant classification other than Delta or Omicron. We removed 20 gene-specific data points with Ct > 40 or Ct < 5 due to possible technical errors. For Ct comparisons, samples with missing data due to failed amplification of a specific gene were removed solely from the analysis of that gene. For the per-affiliation analyses, we removed 6 of 524 (1.1%) BU cases with missing student or employee designations.

### Logistic regression and inference

We fit logistic models on binary variant calls as a function of the date, estimating the proportion of cases that were Omicron over time for each university individually (with data from December 2 – 21), for MA and NE (with data from December 1 – January 1), and for BU by affiliation (student *vs*. employee; with data from December 2 – 21). We documented 95% confidence intervals (CIs) for our model’s parameters, as well as the overdispersion ratio and McFadden’s pseudo-R^2^ (**Supplemental Methods**).

We estimated the date at which the Omicron fraction reached 10%, 50%, and 90%, hereafter defined as O_10_, O_50_, and O_90_. We use the notation ΔO_x, A-B_ = O_x, Population A_ - O_x, Population B_ to represent the difference in the date at which the Omicron fraction reached x% between two populations, and we use the notation ΔO_90-10_ = O_90_ - O_10_ to represent the number of days it took a particular population’s Omicron fraction to rise from 10% to 90%.

We derived point estimates for O_x_ by inverting our regression model, such that:

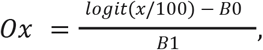

where B_0_ is the model’s intercept and B_1_ is the model’s slope. To generate a standard error for O_x_, we used the delta method^29,30^ with the transformation function *O*_*x*_(x) (as above) and with the mean and covariance of x determined by the coefficients and covariance matrix of our regression model, respectively.

We generated 95% confidence intervals for O_x_ and compared O_x_s between populations by approximating the distribution of O_x_ *via* the family of student’s t distributions (**Supplemental Methods**).

### Case counts

For MA and NE, we summed the confirmed and probable daily cases into the metric total daily cases. We noted weekly variation in case count reporting (*i*.*e*., no MA cases were reported on the weekends; **Supplementary Figure 1**) and thus calculated 7-day rolling averages. We noted smaller-scale variation in case count reporting at IHEs and calculated 3-day rolling averages. To approximate the number of MA cases that were Delta or Omicron, we used our logistic regression model to estimate the Omicron fraction for each day in December 1 – January 15. We scaled case counts by population sizes provided in **Table 1** (IHEs) or in the 2020 census (MA)^31^.

### Ct value comparisons

We compared Ct values for Delta and Omicron cases per institution and per target, given that each university had a unique testing protocol. We compared Ct values for the N1 and N2 genes at BU, the N1 gene at HU, and the N2 and ORF1ab genes at NU. We also compared Ct values at BU per affiliation. We used the Wilcoxon rank-sum test to assess the relationship between SARS-CoV-2 variant (Delta or Omicron) and Ct. We corrected p-values across the comparisons using the Benjamini-Hochberg method.^32^

### Software availability

Custom R scripts are available at https://github.com/bpetros95/omi-uni. De-identified input data is available upon request.

## Results

There was a rapid increase in both daily case count and the Omicron fraction at IHEs in December 2021. In early December, Delta was circulating across Massachusetts and at IHEs, though case rates were higher per-capita in the community outside of IHEs (**Figure 1A**). The Omicron surge at IHEs in mid-December was accompanied by a more modest rise in case counts in MA and NE during the same period, followed by a striking regional surge in late December (**Figure 1A, Supplementary Figure 1**). 1,606 SARS-CoV-2 cases were identified across the 3 institutions (BU, HU, and NU) between December 2 (0% Omicron) and December 21 (91% Omicron). The fraction of cases that were Omicron across the IHEs and NE displayed a classic sigmoid-shaped curve consistent with logistic growth (**Figure 1B, Supplementary Table 2**), moving towards Omicron fixation. Delta diminished in frequency as well as total case count. By January 5, the Harvard University Clinical Laboratory found that 100% of 159 samples tested were Omicron.

**Figure 1.**
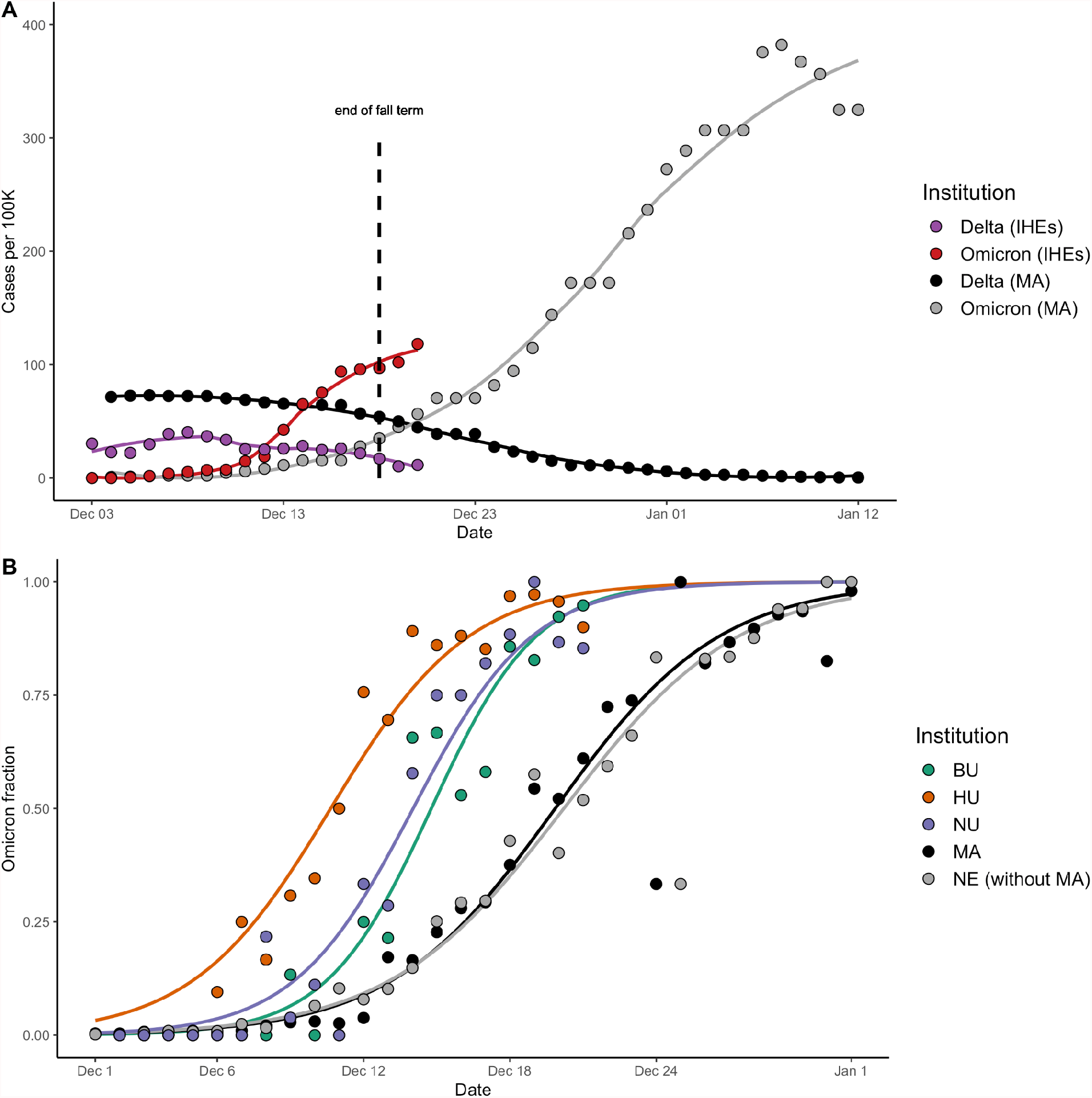
**A**. Cases per 100,000 across the institutes of higher education (IHEs) and MA, stratified by variant. Plotted values are rolling averages over a 3-day (IHEs) or 7-day (MA) window, to account for weekly variation in case reporting. Omicron and Delta variant proportions in MA were inferred from GISAID data (**Methods**). The last day of fall semester finals occurred on Dec 18 (dashed line). Data from Dec 3 – 20 (IHEs) and Dec 4 – Jan 12 (MA). **B**. Proportion of cases that were Omicron, from Dec 2 – 21 (IHEs) and Dec 1 – Jan 1 (MA, NE). Data were modeled using logistic regression. BU, Boston University. HU, Harvard University. NU, Northeastern University. MA, Massachusetts data from GISAID. NE without MA, New England data (excluding the MA data) from GISAID.

Omicron was established earlier and rose to fixation faster at IHEs than in MA as a whole (**Figure 1B**). We noted that MA and NE (without MA) had visually indistinguishable curves, and fitted parameters with highly overlapping confidence intervals (**Supplementary Table 2**). Thus, we compared the timing of Omicron’s trajectory between IHEs and MA, with results generalizable to NE. To compare the timing of Omicron establishment across populations, we generated a metric (O_10_; **Methods**) that estimates the date range at which 10% of the cases were Omicron. O_10_ occurred significantly earlier at IHEs than in MA, by an average of 2.4 days (BU), 3.8 days (NU), and 9.2 days (HU), respectively (**Table 2**; **Supplementary Table 3**). To compare the duration at which Omicron fixated across populations, we generated the metric ΔO_90-10_, the duration (in days) during which Omicron rose from 10% to 90% of cases (**Methods**). ΔO_90-10_ was 9.5 at BU (95% CI, 9.2-9.8), 10.8 at NU (95% CI, 10.4-11.1), 12.5 at HU (95% CI, 12.1-12.9), and 14.8 in MA (95% CI, 14.8-14.9), indicating that the trajectory to Omicron fixation occurred more rapidly at IHEs (**Table 2**). Taken together, these data point towards Omicron’s earlier establishment and faster rise to fixation at IHEs compared to MA or NE.

**Table 2.**
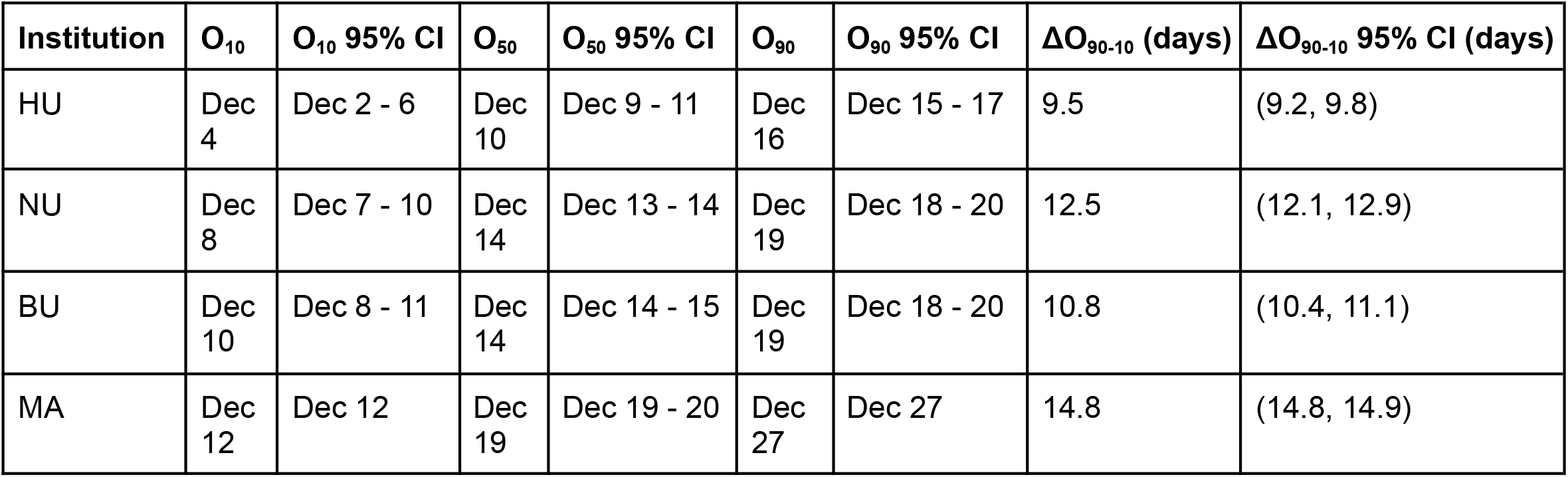
Point estimates and 95% confidence intervals (CIs) for O_x_, the date at which the Omicron fraction equals *x* percent, and for ΔO_90-10_, the duration of time that it takes the Omicron fraction to rise from 10% to 90%. CIs were generated via the student’s t distribution, with estimation of the standard errors via the delta method (**Methods**). BU, Boston University. HU, Harvard University. MA, Massachusetts. NU, Northeastern University.

| Institution | $O_{10}$ | $O_{10}$ 95% CI | $O_{50}$ | $O_{50}$ 95% CI | $O_{90}$ | $O_{90}$ 95% CI | $\Delta O_{90-10}$ (days) | $\Delta O_{90-10}$ 95% CI (days) |
| --- | --- | --- | --- | --- | --- | --- | --- | --- |
| HU | Dec 4 | Dec 2 - 6 | Dec 10 | Dec 9 - 11 | Dec 16 | Dec 15 - 17 | 9.5 | (9.2, 9.8) |
| NU | Dec 8 | Dec 7 - 10 | Dec 14 | Dec 13 - 14 | Dec 19 | Dec 18 - 20 | 12.5 | (12.1, 12.9) |
| BU | Dec 10 | Dec 8 - 11 | Dec 14 | Dec 14 - 15 | Dec 19 | Dec 18 - 20 | 10.8 | (10.4, 11.1) |
| MA | Dec 12 | Dec 12 | Dec 19 | Dec 19 - 20 | Dec 27 | Dec 27 | 14.8 | (14.8, 14.9) |

Next, we found that BU employees displayed Omicron dynamics similar to those of BU students, with a 2-3 day delay in onset. We found no significant association between affiliation (student *vs*. employee) and variant (**Figure 2A**; Fisher’s exact test, p = 0.12, OR = 0.7 with 95% CI 0.5-1.1), with employees accounting for 28.7% (74 Delta, 73 Omicron) of cases and students accounting for 71.3% of cases (157 Delta, 214 Omicron; **Figure 2B**). We again used O_10_ (**Methods**) to compare the timing of Omicron establishment between populations. O_10_ occurred significantly earlier among BU students relative to BU employees (by an average of 2.8 days) and MA (by an average of 3.0 days); **Figure 2C**; **Table 3**; **Supplementary Table 5**). We compared ΔO_90-10_ as a metric of time to fixation, which was 8.5 among BU employees (95% CI, 7.9-9.1) and 9.5 among BU students (95% CI, 9.1-9.9; **Table 3**). ΔO_90-10_ was more comparable between BU students and employees than BU was to other IHEs (**Table 2**), and markedly different in MA (14.8, 95% CI, 14.8-14.9), indicating that employees’ trajectory resembles that of students (**Table 3**). Taken together, we found that employees and students have parallel Omicron trajectories, with a lag time between two and three days.

**Table 3.** Point estimates and 95% confidence intervals (CIs) for O_x_, the date at which the Omicron fraction equals *x* percent, and for ΔO_90-10_, the duration of time that it takes the Omicron fraction to rise from 10% to 90%. CIs were generated via the student’s t distribution, with estimation of the standard errors via the delta method (**Methods**). BU, Boston University. MA, Massachusetts.

| **Affiliation** | **O<sub>10</sub>** | **O<sub>10</sub> 95% CI** | **O<sub>50</sub>** | **O<sub>50</sub> 95% CI** | **O<sub>90</sub>** | **O<sub>90</sub> 95% CI** | **$\Delta O_{90-10}$ (days)** | **$\Delta O_{90-10}$ 95% CI (days)** |
| --- | --- | --- | --- | --- | --- | --- | --- | --- |
| BU employees | Dec 12 | Dec 10 - 14 | Dec 16 | Dec 15 - 17 | Dec 20 | Dec 19 - 22 | 8.5 | (7.9, 9.1) |
| BU students | Dec 9 | Dec 7 - 10 | Dec 14 | Dec 13 - 14 | Dec 18 | Dec 17 - 20 | 9.5 | (9.1, 9.9) |
| MA | Dec 12 | Dec 12 | Dec 19 | Dec 19 - 20 | Dec 27 | Dec 27 | 14.8 | (14.8, 14.9) |

**Figure 2.**
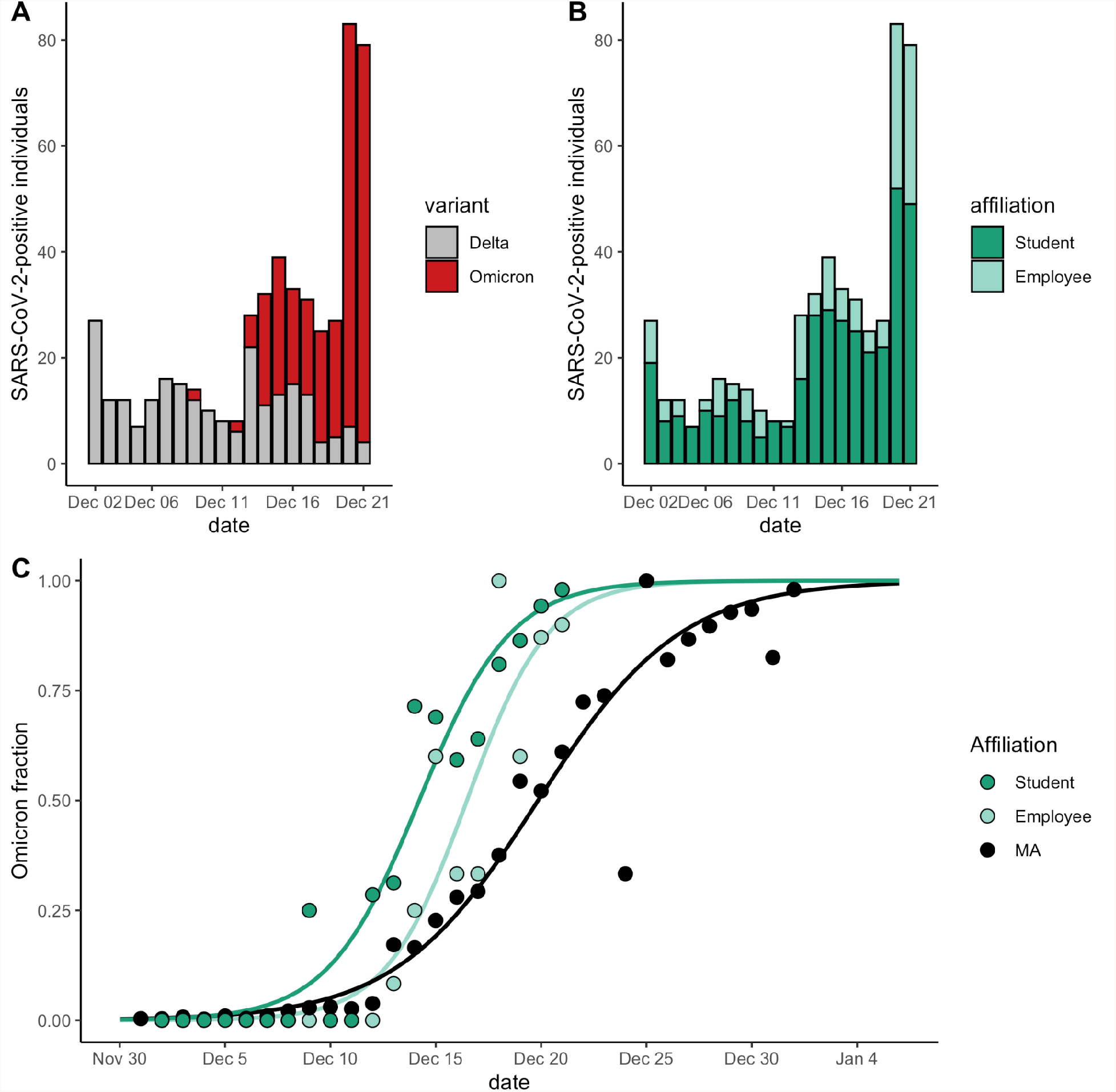
**A**. Total number of cases at BU, stratified by variant and stacked, from December 2-21. Gray, Delta. Red, Omicron. **B**. Total number of cases at BU, stratified by affiliation status and stacked, from December 2-21. Light green, employees. Green, students. **C**. Proportion of cases that were Omicron, from Dec 2 – 21 (BU students and employees) and Dec 1 – Jan 1 (MA). Data were modeled using logistic regression. BU, Boston University. MA, Massachusetts data from GISAID.

Finally, we compared Ct values across variants, and found that Omicron samples did not have lower Cts (*i*.*e*., higher viral loads) than Delta samples, suggesting that increased Omicron transmission is not driven by higher viral loads (**Figure 3**). At BU and HU, N1-gene Ct values were significantly higher, by an average of 2.2 (p = 0.0002) and 3.1 (p < 0.0001) respectively, in Omicron *vs*. Delta samples (**Figure 3AB**; **Supplementary Table 6**). This trend was recapitulated for N2-gene Ct values at BU (Omicron Cts an average of 2.0 higher, p = 0.0007; **Figure 3C**; **Supplementary Table 6**). At NU, neither the N2-nor the ORF1ab-gene Cts differed by variant (**Figure 3D**; **Supplementary Figure 2**; **Supplementary Table 6**). We found no difference in BU’s Ct values by affiliation status (*i*.*e*., student *vs*. employee; N1 gene, p = 0.91; N2 gene, p = 0.81), and that Ct-by-variant trends identified at BU are conserved when conditioning the data on an affiliation (**Supplementary Figure 3**; **Supplementary Table 7**). In summary, we found that, despite differences in testing cadence, testing platform, and demographics among IHEs, Omicron viral Cts were always higher or indistinguishable from Delta Cts.

**Figure 3.**
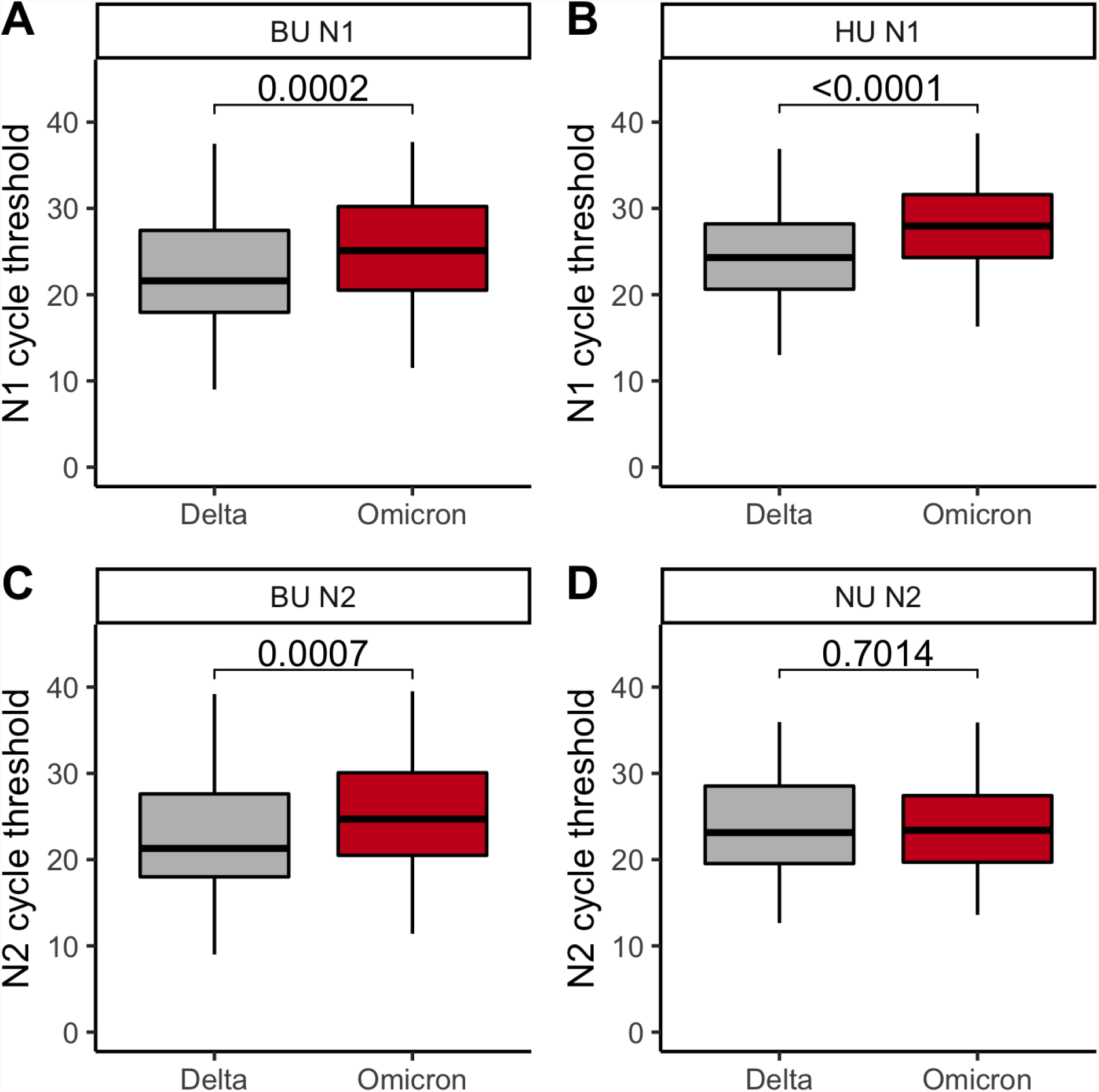
**AB**. N1 cycle threshold for Delta *vs*. Omicron cases at BU (**A**) and HU (**B**). **CD**. N2 cycle threshold for Delta *vs*. Omicron cases at BU (**C**) and NU (**D**). **ABCD**. Gray, Delta. Red, Omicron. The first, second, and third quartiles are within the box, with the median line bolded. The whisker length is 1.5 times the interquartile range (IQR), unless the furthest point is less than 1.5*(IQR) from the quartile. Outliers are displayed as points. P-values via Wilcoxon rank sum test and corrected via Benjamini-Hochberg method (across the 4 comparisons of **Figure 3** and the 1 comparison of **Supplementary Figure 2**).

## Discussion

Here, we document Omicron’s swift spread through Boston-based IHEs in December 2021, leading to unprecedented increases in daily case counts. Though all three IHEs and the urban environment in which they are located were experiencing Delta transmission at the time of Omicron introduction, Omicron rapidly became the dominant variant. Over an 8-13 day period, variant proportions converted from > 90% Delta to > 90% Omicron. Importantly, the rapid increase in Omicron case counts was identified in highly vaccinated populations in which Omicron’s viral load, as inferred from anterior nares diagnostic Ct, was comparable to or lower than that of Delta, consistent with other reports^33,34^. This highlights that Omicron’s fitness is neither driven by a higher viral load nor reliant on an immunologically naive population.

Though the date of establishment differed at the three IHEs, the dynamics of Omicron takeover were strikingly similar. The rapid rise in the Omicron fraction was offset by 1-4 days between universities, though we cannot rule out the differences in testing cadence as the cause of the lag. However, the dynamics of Omicron dominance were similar across campuses despite differences in testing programs, on-campus *vs*. off-campus housing, and variant designation technologies. Additionally, the time to fixation for BU employees was more comparable to that of BU students than that of the state, supporting a transmission mode that is independent of the residential nature of college campuses.

In contrast to the early establishment and dominance in the IHEs in our study, Omicron’s procession towards fixation in Massachusetts occurred more slowly. While some of the difference in introduction time could be accounted for by earlier detection of cases in asymptomatic testing programs, the differences in slope and time to fixation cannot be explained by this factor. These dynamics are consistent with the classic dynamics of overdispersion in transmission, in which clusters of cases are responsible for the majority of spread, and the early stages of establishment within a community are stochastic and scale with the number of introductions^35,36^. Overdispersion in SARS-CoV-2 transmission is well documented^35,37–39^ and our work is consistent with the continuation of this phenomenon with Omicron. IHEs are not siloed in their interaction networks; however, the proportion of interactions within an IHE’s network is greater than those that exit into the community, and as a result clusters of transmission are readily detected via robust screening. The ability of Omicron to rapidly spread through a vaccinated population means that even communities that appear to escape the initial peak of Omicron in the US require continued caution. For example, rural communities may experience late introductions of Omicron, and may not notice its arrival until a significant proportion of the population has become infected. This is of particular concern in areas with low vaccination rates, where a rapid rise in case counts can more readily overwhelm health care systems.

There are several limitations to the generalizability of this study. While universities contain individuals from different communities, features of a university such as the age distribution, the availability of residential life, and extracurricular activities could influence Omicron dynamics. Furthermore, the degree to which potential superspreader events could be important – not just for the initial seeding of Omicron, but for its continued spread – is not captured here. For example, it is possible that the spread of Omicron in other types of communities may be slower if the structure of the social network differs^40^, resulting in fewer opportunities for clustered transmission. Additionally, our work is limited by a few differences between MA and IHEs. For example, reduced vaccination rates in MA, relative to IHEs, may contribute to the relative fitness of Delta *vs*. Omicron. Moreover, MA testing may be biased towards Delta samples if symptomatic testing occurs more frequently with Delta than with Omicron.

What can we learn from the spread of Omicron through universities that could help us to mitigate future waves of SARS-CoV-2 or other pathogens? First, for SARS-CoV-2, sites that have characteristics like IHEs can be informative early detection sites. We note two of many reasons: (1) IHEs contain individuals from a variety of backgrounds who travel and intermix at the university and in the larger community, and (2) IHEs have implemented university-wide asymptomatic screening programs. Screening programs like these could be used to catch and categorize infections well before trends are noted in the larger community, and have the potential to forecast anticipated testing needs and hospital admissions. Second, it is extremely difficult to stop the spread of a highly transmissible virus once it has become established in a community. BU, HU, and NU controlled the spread of previous variants of SARS-CoV-2 *via* a combination of high-cadence testing, isolation of positive individuals, contract tracing, quarantining of close contacts, social distancing, masking, vaccination requirements, and ventilation improvements. These measures were not sufficient to stop the spread of Omicron, emphasizing the need for continued surveillance programs to rapidly identify and mitigate outbreaks before they become pandemics.

## Author Contributions

B.A.P and J.T. accessed and verified the underlying data reported in the manuscript. B.A.P. conducted statistical analyses under the guidance of L.F.W. and E.D.K. B.A.P. and J.T. produced software and visualizations. J.T. collected and curated Boston University data with help from L.D.-S., J.T.P., K.R.J., T.B., L.L., C.M.K., and D.H.H. N.L.W. collected and curated mCARMEN data with help from M.R.B., S.T.D., and T.G.N. Variant-specific PCR was collected and curated by M.C. and M.H. using custom primers and probes designed by M.S., M.G., D.T., and E.W. P.N. aided with project administration. B.P.T. curated CDC and GISAID data. J.R.A. and S.Y. collected and curated Northeastern University data. B.A.P. wrote the original draft of the manuscript with feedback provided by W.P.H., P.C.S., J.H.C., and M.S. N.L.W., P.N., K.R.J., T.B., W.H., P.C.S., J.H.C., and M.S. reviewed and edited the manuscript. W.P.H., P.C.S., J.H.C., and M.S. conceptualized and supervised the study, and reviewed and edited the final manuscript.

## Acknowledgements

We gratefully acknowledge the students and employees at each participating university, and the staff at the BU, HU, and NU clinical testing laboratories. We gratefully acknowledge the authors from the originating laboratories responsible for obtaining the specimens and the submitting laboratories where genetic sequence data were generated and shared *via* the GISAID Initiative, on which this research is based. All submitters of data may be contacted directly *via* www.gisaid.org. Access to patient sample metadata was facilitated by the Massachusetts Consortium on Pathogen Readiness (MassCPR).

## Funding

B.A.P. is supported by the National Institute of General Medical Sciences (grant T32GM007753). L.F.W. is supported by the National Institute of General Medical Sciences (grant R35GM141821). J.H.C. acknowledges funding from Boston University for SARS-CoV-2 surveillance, from the Massachusetts Consortium on Pathogen Readiness (MassCPR), and from the China Evergrande Group. W.P.H., T.P.B., and K.R.J. also acknowledge support from MassCPR. T.B. is supported by the National Institute of Health (NIAID K23 grant AI152930-01A1). B.L.M. is supported by the Centers for Disease Control and Prevention SARS-CoV-2 Baseline Genomic Surveillance contract (75D30121C10501 to Broad Institute via the Clinical Research Sequencing Platform, LLC), a CDC Broad Agency Announcement (75D30120C09605 to B.L.M.), and the Rockefeller Foundation (2021 HTH 013 to B.L.M. and P.C.S.). M.S. is supported by the National Institute of Health (grant 5R01GM120122). The content is solely the responsibility of the authors and does not necessarily represent the official views of the National Institute of General Medical Sciences or the National Institutes of Health.

## Conflicts of Interest

M.S. is co-founder of, shareholder in, and advisor to Rhinostics Inc. M.G, D.T., S.R., and E.W. are employed by Integrated DNA Technologies. C.M.K. is co-founder of Biosens8, Inc. J.H.C. is a consultant for Cell Signaling Technologies. W.P.H. serves on Scientific Advisory Boards for Biobot Analytics Inc. and Merck. P.C.S. is a founder of and shareholder in Sherlock Biosciences, and is both on the Board and serves as shareholder in the Danaher Corporation.

## Supplemental Methods

### Experimental methods

At BU, affiliates self-collected anterior nares specimens, which were analyzed by RT-qPCR as previously described^11^. Variant status was assessed using amplicon-based viral sequencing^22^ with the ARTIC v4 primer set and the PANGOLIN lineage classification algorithm, as previously described.

At HU, affiliates self-collected anterior nares specimens, which were rehydrated with 300 uL phosphate-buffered saline (PBS) and inactivated at 65 degrees Celsius. RT-qPCR was performed using the Quaeris SARS-CoV-2 assay^20^, and Cts for the SARS-CoV-2 N1 and RdRP genes (and the human RNase P gene, as a positive control) were determined using the Applied Biosystems QuantStudio 7 Real-Time PCR instrument (software version 1.7).

The mCARMEN platform^23^ distinguishes between the Delta and Omicron variants using Spike gene mutation signatures (Delta = del156/157, L452R; Omicron = del69/70, K417N, S477N, N501Y, P681H). It was run on 101 samples with 97% concordance to next-generation sequencing (NGS)^23^ and on 1,557 samples with 99.5% concordance to NGS (ref. 23, update in prep). RNA extraction was performed using the Thermo Fisher Scientific Applied Biosystems™ MagMAX™ mirVana Total RNA Isolation Kit and the Spike gene was amplified using the Thermo Fisher SuperScript™ IV One-Step RT-PCR System prior to running mCARMEN.

The variant-specific PCR assay discriminates between Delta and Omicron *via* detection of the following Spike protein SNVs: L452R, Q498R, N501Y (primer and probe sequences: **Supplementary Table 1**). Primers and probes were verified on samples that were confirmed, *via* genomic sequencing, to be Delta or Omicron. Of the 384 samples tested, 343 yielded a call in 2 or 3 of the variant PCR reactions, leading to a variant designation; the 41 samples with only a single variant PCR call were excluded from the data set.

At Northeastern University, affiliates self-collected anterior nares specimens, and RNA was extracted from the clinical specimens using the Thermo Fisher Scientific Applied Biosystems™ MagMAX™ Viral/Pathogen II (MVP II) Nucleic Acid Isolation Kit. Cycle thresholds were determined *via* RT-qPCR, conducted with the Thermo Fisher Scientific Applied Biosystems™ TaqPath™ COVID-19 Combo Kit (with primers and probes specific to the N2, ORF1ab, and S genes^21^) according to the manufacturer’s instructions. Samples with S-gene target failure (SGTF) were designated as Omicron and samples with S-gene target amplification were designated as Delta.

### Analytic methods Software specifications

Analyses were run in R version 4.0.2 (2020-06-22) on a 64-bit Linux/GNU PC and on a Macbook Pro with a Darwin 17.0 platform and R version 4.1.1 (2021-08-10), with no issue reproducing analyses on each machine. Required packages include base, boot, cowplot, ggpubr, msm, reshape2, rstatix, stats, tidyverse, and zoo.

### Logistic regression

We downloaded cases counts by state over time from the CDC^25^ on January 19, 2022. We downloaded MA and NE GISAID data^26–28^ on January 12, 2022. We noted that MA was overrepresented in the NE data (MA contains approximately 47% of the region’s population, but 72% of available sequences), and thus removed MA from the NE data.

We fit logistic curves to the data using R’s generalized linear models package:

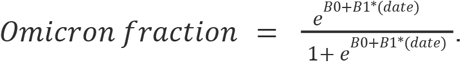

We calculated overdispersion ratios^41,42^ to assess for the possibility of unaccounted-for variability in our models:

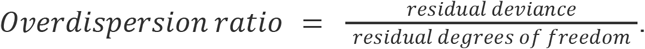

We also calculated McFadden’s^43,44^ pseudo-R^2^ as follows, where *L* is the likelihood function, the null model (“null”) regresses our data as a function of a constant (*i*.*e*., 1), and the alternative model (“model”) regresses our data as a function of the date:

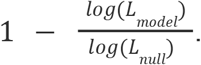

The logistic growth model was a reasonable choice: (1) McFadden’s pseudo-R^2^ was in [0.31, 0.56] for all seven regression models (**Supplementary Tables 2 & 4**); and (2) there was no evidence of overdispersion, with ratios in [0.56 - 0.81] (**Supplementary Tables 2 & 4**).

We generated standard errors for O_x_ using the delta method (from R’s multi-state Markov package), and used these standard errors for inference as follows. In all cases, the sample size for each population was the number of days in our logistic regression models (*e*.*g*., for BU, sample size = 20 as we included data from December 2-21):

1. We generated 95% CIs for O_x_ by assuming O_x_ approximately follows a student’s t distribution (*i*.*e*., point estimate +/- *t*\*(standard error)), where *t* is the 97.5^th^ percentile of a student’s t distribution with degrees of freedom equal to one less than the sample size.
2. We compared O_x_ between populations – *i*.*e*., ΔO_x,A-B_ = O_x, Population A_ - O_x, Population B_ – by running a student’s t-test.
3. We generated 95% CIs for ΔO_90-10_ = O_90_ - O_10_ by assuming that ΔO_90-10_ approximately follows a student’s t distribution (*i*.*e*., point estimate +/- *t*\*(standard error)), where *t* is the 97.5^th^ percentile of a student’s t distribution with degrees of freedom equal to one less than the sample size.

### Comparisons of logistic fits

The logistic regression models were also compared, with the intercept parameter providing a metric for Omicron introduction time, and the slope parameter serving as a metric of the speed of fixation. The intercept parameters of our logistic regression models (B_0_) were significantly lower at BU and at NU than in MA or NE, and trends earlier at HU than in MA or NE (**Supplementary Table 2**). Moreover, the slope parameters of our logistic regression models (B_1_) were significantly steeper at BU (95% CI, 0.39-0.55) and NU (95% CI, 0.34-0.48) than in MA or NE (95% CI, 0.29-0.30), and trends steeper at HU (95% CI, 0.29-0.41) than in MA or NE (**Supplementary Table 2**). Taken together, our models provide support for earlier introductions and increased speed to fixation at IHEs.

We compared our intercepts and slopes across BU affiliations as well. The intercept parameters of our logistic regression models (B_0_) were significantly lower among both BU students and employees than in MA (**Supplementary Table 5**). Moreover, the slope parameters of our logistic regression models (B_1_) were significantly steeper among BU employees (95% CI, 0.37-0.70) and students (95% CI, 0.37-0.57) than in MA (95% CI, 0.29-0.30; **Supplementary Table 5**). These data suggest that BU employees were similar to students with respect to their trajectories to Omicron fixation.

**Supplementary Table 1.**
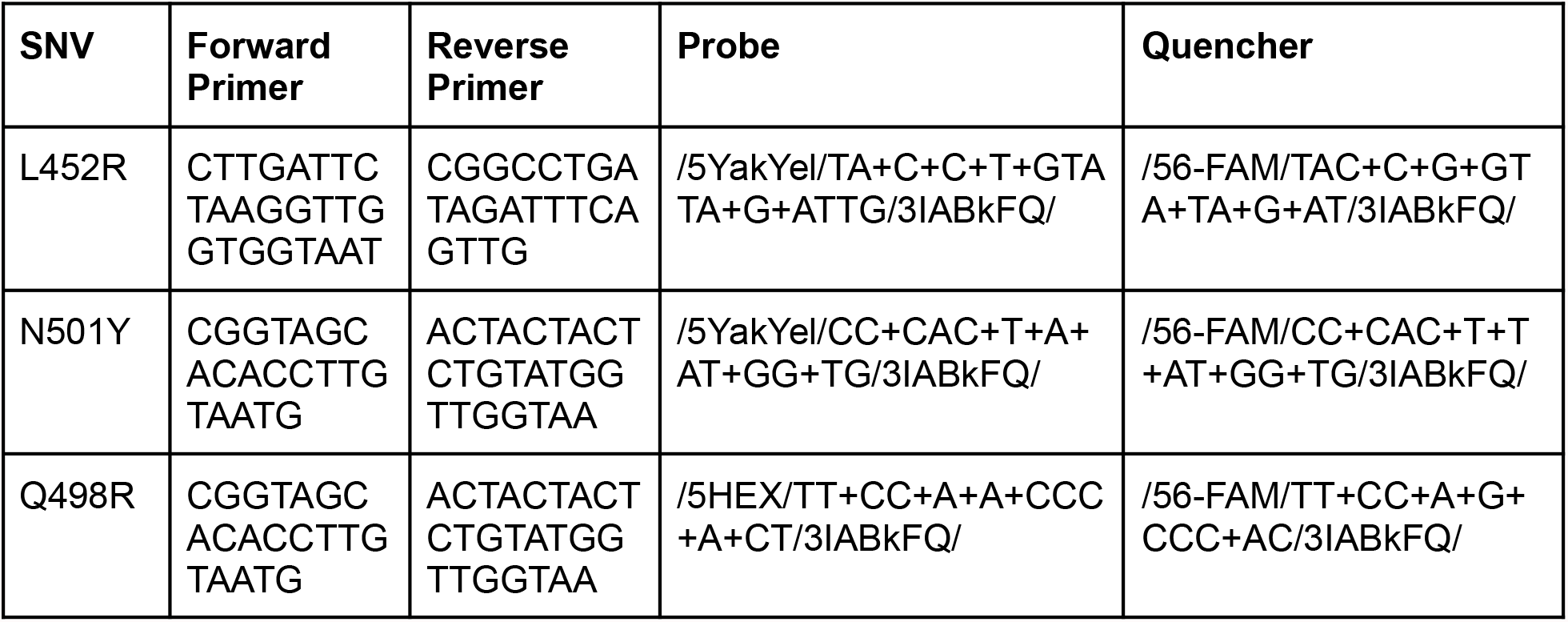
Primer and probe sequences used to determine whether a subset of samples at Harvard University were Delta *vs*. Omicron.

**Supplementary Figure 1.**
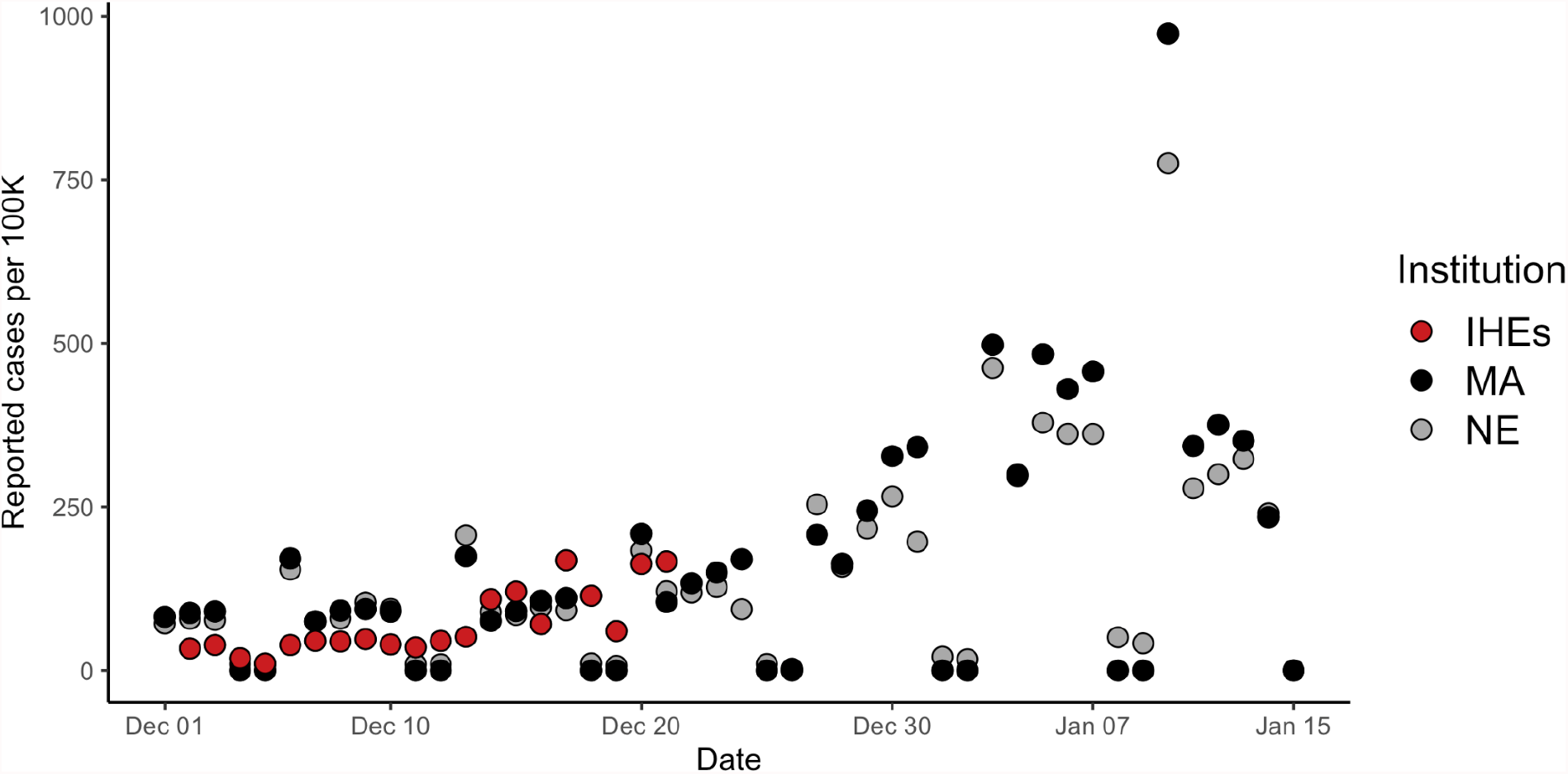
Reported daily cases per 100,000 across the institutes of higher education (IHEs), Massachusetts (MA), and NE (New England). No methods were used to account for weekly variation in case reporting. Data from Dec 2 – 21 (IHEs) and Dec 1 – Jan 15 (MA, NE).

**Supplementary Table 2.** Logistic regression point estimates and 95% confidence intervals (CI) for intercept (B_0_) and slope (B_1_) parameters for each university. * signifies that the 95% confidence interval does not include 0. BU, Boston University. HU, Harvard University. MA, Massachusetts. NU, Northeastern University. NE, New England.

| Institution | $B_0$ | $B_0$ 95% CI | $B_1$ | $B_1$ 95% CI | McFadden's $R^2$ | Overdispersion ratio |
| --- | --- | --- | --- | --- | --- | --- |
| BU | -8787.44 | (-10408.66, -7358.17)* | 0.46 | (0.39, 0.55)* | 0.47 | 0.73 |
| HU | -6672.11 | (-7847.37, -5595.88)* | 0.35 | (0.29, 0.41)* | 0.31 | 0.78 |
| NU | -7751.57 | (-9194.38, -6462.90)* | 0.41 | (0.34, 0.48)* | 0.41 | 0.81 |
| MA | -5622.16 | (-5769.38, -5478.22)* | 0.29 | (0.29, 0.30)* | 0.56 | 0.56 |
| NE | -5283.40 | (-5524.90, -5049.55)* | 0.30 | (0.29, 0.30)* | 0.45 | 0.58 |

**Supplementary Table 3.** Point estimates and p-values for ΔO_x,A-B_, the difference, in days, between O_x, Institution A_ and O_x, Institution B_. P-values were generated via the student’s two-sample t test and corrected via the Benjamini-Hochberg method. BU, Boston University. HU, Harvard University. MA, Massachusetts. NU, Northeastern University.

| Institution A | Institution B | x | $\Delta O_{x,A-B}$ (days) | P-value |
| --- | --- | --- | --- | --- |
| BU | HU | 0.1 | 5.6 | < 0.0001 |
| BU | MA | 0.1 | 2.4 | < 0.0001 |
| BU | NU | 0.1 | 1.4 | < 0.0001 |
| HU | MA | 0.1 | 8.0 | < 0.0001 |
| HU | NU | 0.1 | -4.3 | < 0.0001 |
| MA | NU | 0.1 | 3.8 | < 0.0001 |
| BU | HU | 0.5 | 4.1 | < 0.0001 |
| BU | MA | 0.5 | 5.1 | < 0.0001 |
| BU | NU | 0.5 | 0.7 | < 0.0001 |
| HU | MA | 0.5 | 9.2 | < 0.0001 |
| HU | NU | 0.5 | -3.4 | < 0.0001 |
| MA | NU | 0.5 | 5.8 | < 0.0001 |
| BU | HU | 0.9 | 2.6 | < 0.0001 |
| BU | MA | 0.9 | 7.8 | < 0.0001 |
| BU | NU | 0.9 | 0.1 | 0.44 |
| HU | MA | 0.9 | 10.4 | < 0.0001 |
| HU | NU | 0.9 | -2.5 | < 0.0001 |
| MA | NU | 0.9 | 7.9 | < 0.0001 |

**Supplementary Table 4.**
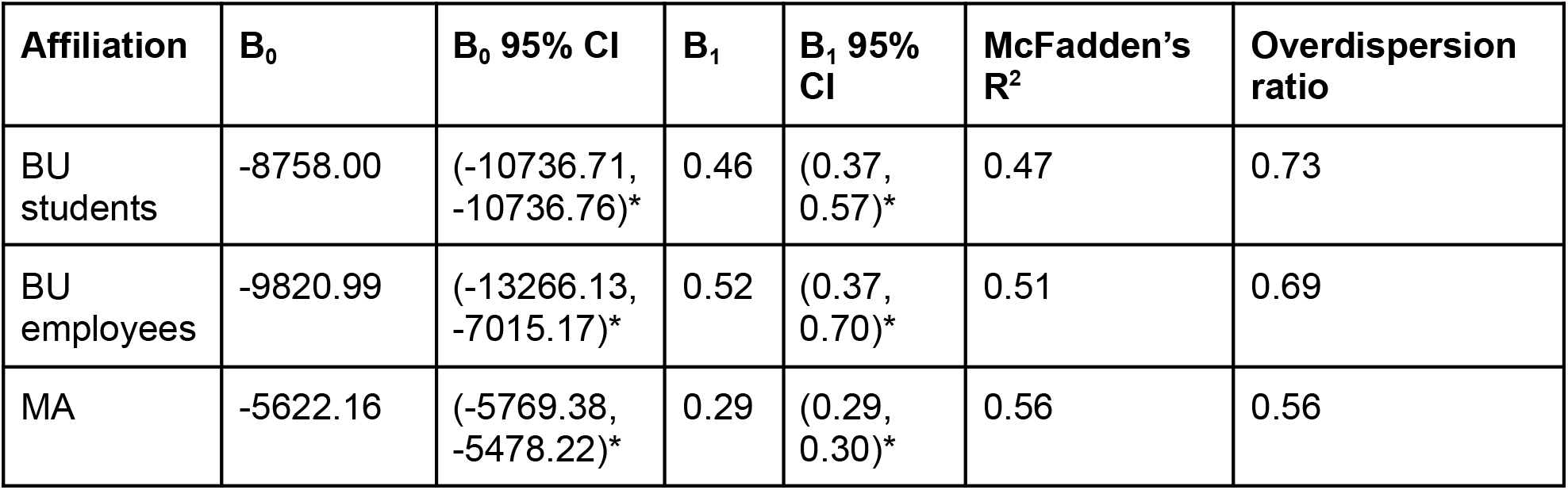
Logistic regression point estimates and 95% confidence intervals (CI) for intercept (B_0_) and slope (B_1_) parameters for students and for employees at Boston University (BU). * signifies that the 95% confidence interval does not include 0. MA, Massachusetts.

**Supplementary Table 5.**
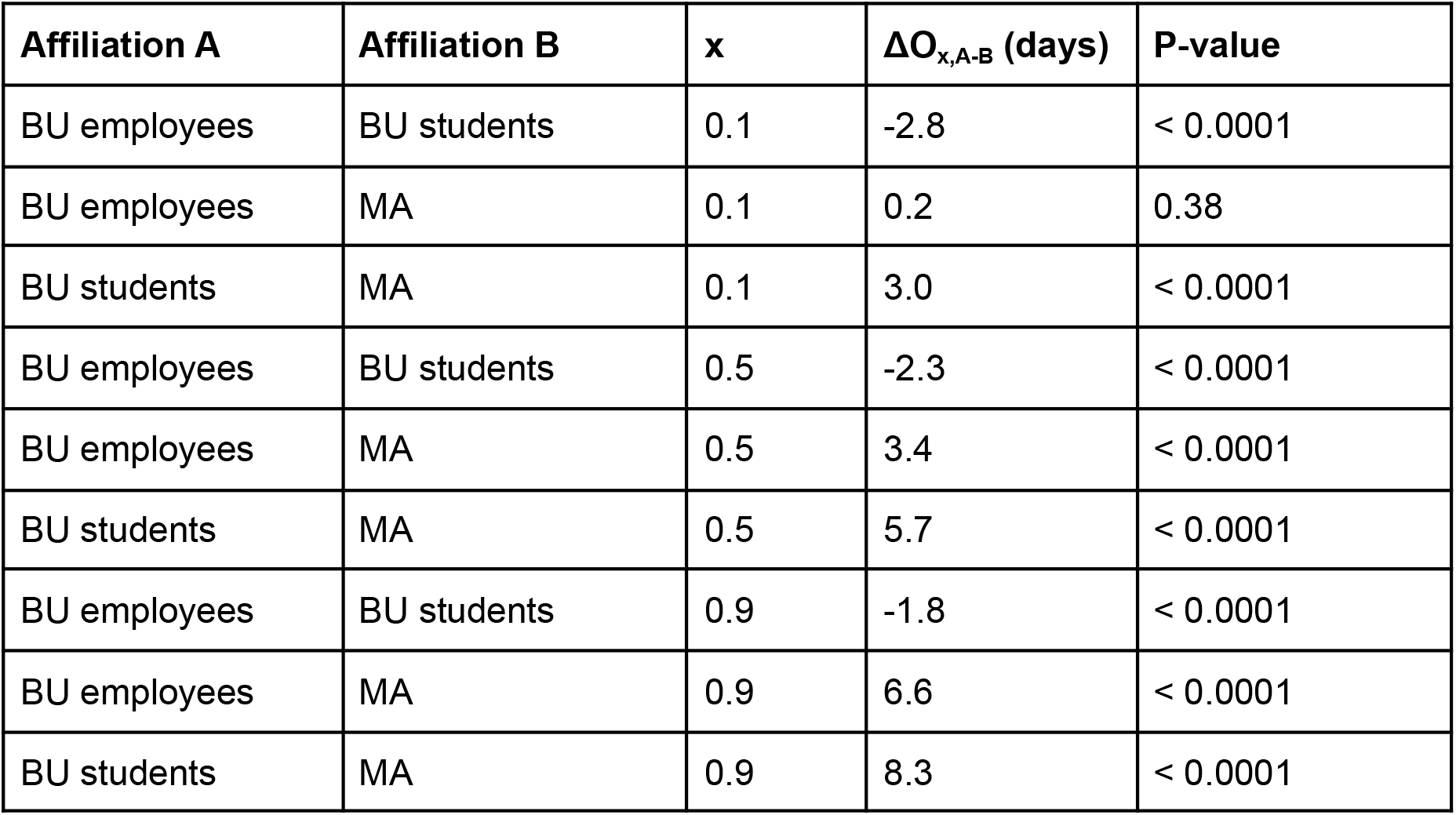
Point estimates and p-values for ΔO_x,A-B_, the difference, in days, between O_x, Affiliation A_ and O_x, Affiliation B_. P-values were generated via the student’s two-sample t test and corrected via the Benjamini-Hochberg method. BU, Boston University. MA, Massachusetts.

**Supplementary Figure 2.**
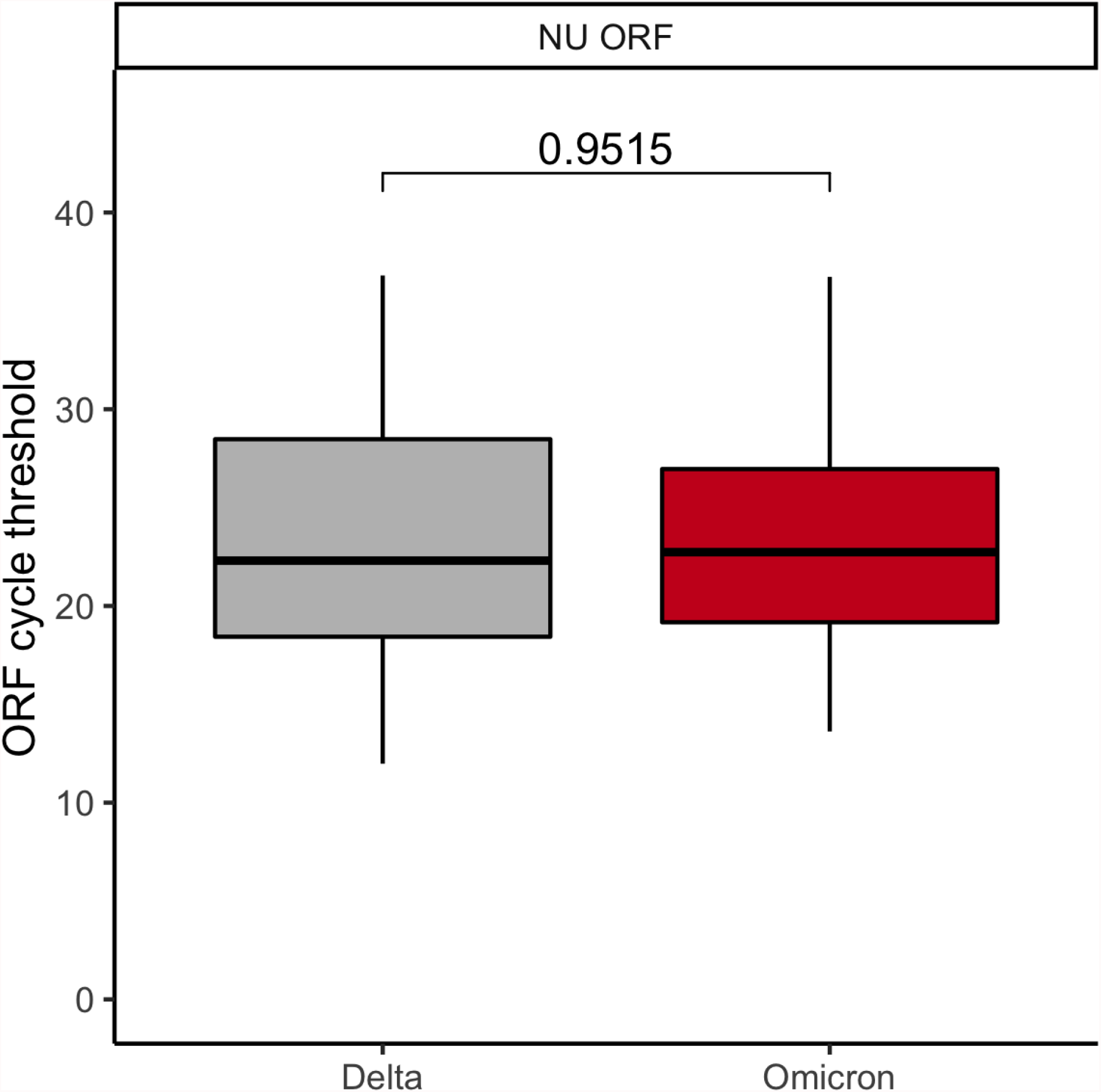
ORF1ab cycle threshold (Ct) for Delta *vs*. Omicron cases at NU. Gray, Delta. Red, Omicron. The first, second, and third quartiles are within the box, with the median line bolded. The whisker length is 1.5 times the interquartile range (IQR), unless the furthest point is less than 1.5*(IQR) from the quartile. Outliers are displayed as points. P-value via Wilcoxon rank sum test and corrected via Benjamini-Hochberg method (across the 4 comparisons of **Figure 3** and the 1 comparison of **Supplementary Figure 2**).

**Supplementary Table 6.** Mean and median cycle threshold (Ct) values, per institution and per variant. BU, Boston University. HU, Harvard University. NU, Northeastern University.

| **Institution** | **Variant** | **Target** | **Median** | **Mean** |
| --- | --- | --- | --- | --- |
| BU | Delta | N1 | 21.6 | 22.9 |
| BU | Omicron | N1 | 25.1 | 25.2 |
| BU | Delta | N2 | 21.3 | 22.9 |
| BU | Omicron | N2 | 24.7 | 24.9 |
| HU | Delta | N1 | 24.3 | 24.6 |
| HU | Omicron | N1 | 28.0 | 27.7 |
| NU | Delta | N2 | 23.1 | 23.9 |
| NU | Omicron | N2 | 23.4 | 23.6 |
| NU | Delta | ORF | 22.3 | 23.4 |
| NU | Omicron | ORF | 22.7 | 23.1 |

**Supplementary Figure 3.**
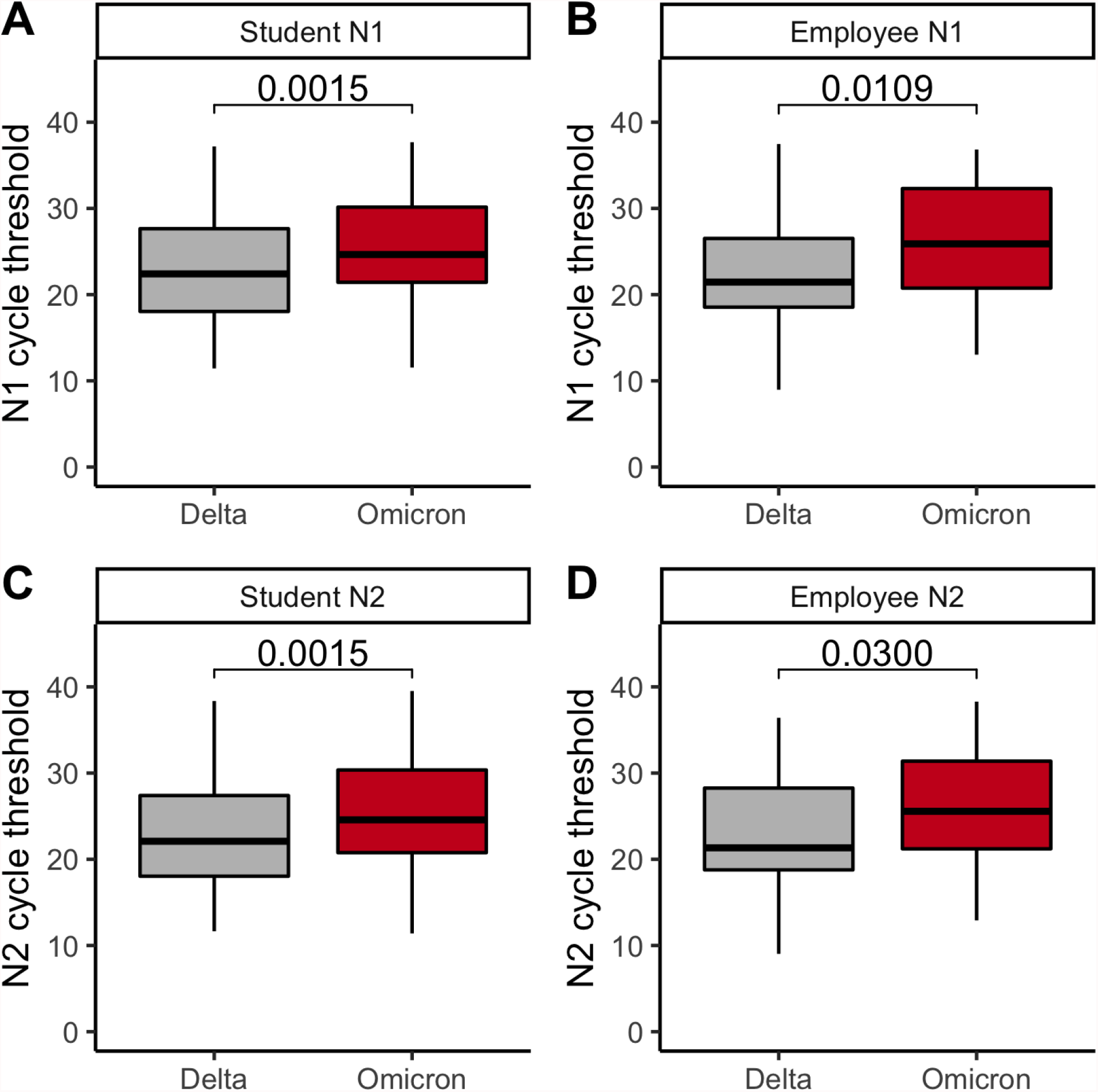
**AB**. N1 cycle threshold for Delta *vs*. Omicron cases among BU students (**A**) and employees (**B**). **CD**. N2 cycle threshold for Delta *vs*. Omicron cases among BU’s students (**C**) and employees (**D**). **ABCD**. Gray, Delta. Red, Omicron. The first, second, and third quartiles are within the box, with the median line bolded. The whisker length is 1.5 times the interquartile range (IQR), unless the furthest point is less than 1.5*(IQR) from the quartile. Outliers are displayed as points. P-values via Wilcoxon rank sum test and corrected via Benjamini-Hochberg method (across the 4 comparisons of **Supplementary Figure 3**). BU, Boston University.

**Supplementary Table 7.**
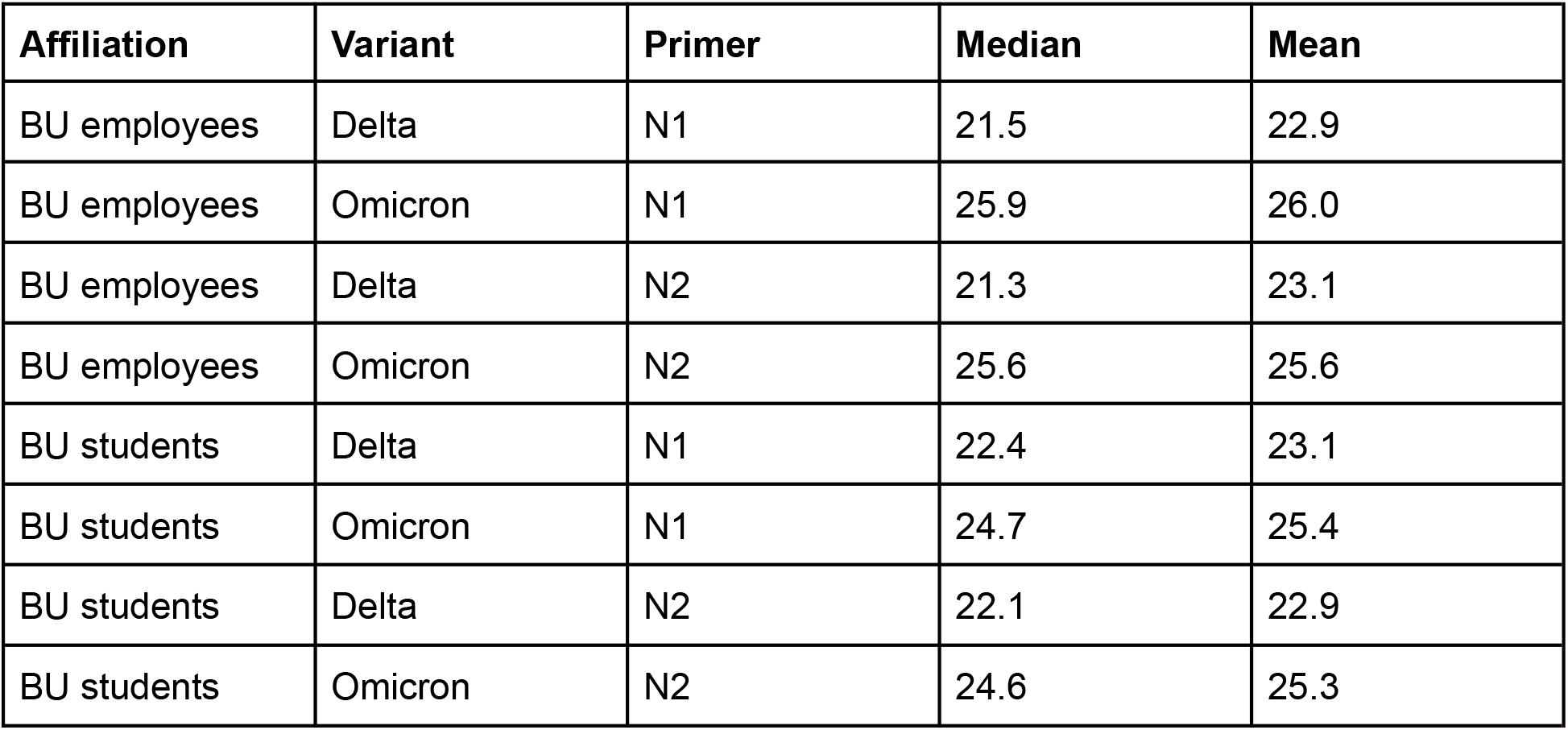
Mean and median cycle threshold (Ct) values at BU, per affiliation and per variant. BU, Boston University.

| **Affiliation** | **Variant** | **Primer** | **Median** | **Mean** |
| --- | --- | --- | --- | --- |
| BU employees | Delta | N1 | 21.5 | 22.9 |
| BU employees | Omicron | N1 | 25.9 | 26.0 |
| BU employees | Delta | N2 | 21.3 | 23.1 |
| BU employees | Omicron | N2 | 25.6 | 25.6 |
| BU students | Delta | N1 | 22.4 | 23.1 |
| BU students | Omicron | N1 | 24.7 | 25.4 |
| BU students | Delta | N2 | 22.1 | 22.9 |
| BU students | Omicron | N2 | 24.6 | 25.3 |

## Notes

### Author Declarations

Harvard Longwood Campus IRB (protocol #20-1877) gave ethical approval for work with Harvard University samples. These samples were covered by an exempt determination (EX-7295) at the Broad Institute of MIT and Harvard. Boston University IRB (protocol #6122E) gave ethical approval for work with Boston University samples. Northeastern University IRB (protocol #21-02-07) provided an exempt determination for Northeastern University samples, whose metadata was shared with the authors under Data Use Agreement 20-1481.

